# Familial risk of myocardial infarction with non-obstructive and obstructive coronary arteries-A nation-wide cohort study

**DOI:** 10.1101/2024.01.29.24301968

**Authors:** Felicia HK Hakansson, Per Svensson, Hans J Pettersson, Ewa Ehrenborg, Jonas Spaak, Anna M Nordenskjold, Kai M Eggers, Per Tornvall

**Affiliations:** Department of Clinical Science and Education Södersjukhuset, Karolinska Institutet, Stockholm, Sweden; Division of Cardiovascular Medicine, Department of Medicine Solna, Karolinska Institutet, Stockholm, Sweden; Department of Cardiology and Clinical Science and Education Danderyds Hospital, Karolinska Institutet, Stockholm, Sweden; Department of Cardiology, Faculty of Medicine and Health, Örebro University, Örebro, Sweden; Department of Medical Sciences, Cardiology, Uppsala University, Uppsala, Sweden; Department of Cardiology Södersjukhuset, Stockholm, Sweden

**Keywords:** MINOCA, MI-CAD, familial risk, coronary angiography

## Abstract

**Background:** The familial risk among patients with myocardial infarction with non-obstructive coronary arteries (MINOCA) is unknown. Previous studies of family history in myocardial infarction (MI), have not made a distinction between MINOCA and MI due to coronary artery disease (MI-CAD), based on angiographic findings.

**Objectives:** To investigate familial risk of MI without and with obstructive coronary arteries.

**Methods:** A register-based cohort study with a total of 15,462 MINOCA cases, 204,424 MI-CAD cases, 38,220 control subjects without MI and with non-obstructive coronary arteries. First-degree relatives were identified 1995-2020. Cox proportional hazard regression models were used to compare familial risk in MINOCA and MI-CAD with control subjects.

**Results:** During a mean follow-up of 8.1 ± 4.2 years, MINOCA occurred in 1.0% of first-degree relatives with MINOCA whereas MI-CAD occurred in 9.7% of first-degree relatives of MINOCA. The age- and sex-adjusted hazard ratio (HR) for a MINOCA-relative experiencing MINOCA and MI-CAD, compared to control subjects, was 0.99 (95% confidence interval [CI] 0.80-1.23) and 1.10 (95% CI 1.03-1.18), respectively. During a mean follow-up of 8.5 ±4.8 years, MI-CAD occurred in 12.2% of first-degree relatives with MI-CAD with age- and sex-adjusted HR 1.43 (95% CI 1.37-1.49).

**Conclusions:** No increased familial risk of MINOCA was observed for MINOCA-patients whereas there was an increased familial risk for MI-CAD when compared to control subjects. These results may indicate that genetic factors and shared environmental factors within a family leading to CAD are important also for MINOCA, thus MI-CAD and MINOCA could share underlying mechanisms.

**Condensed Abstract:** In this register-based nation-wide cohort study including 15,462 MINOCA cases, 204,424 MI-CAD cases, and 38,220 control subjects, studied between 1995-2020, we show that there is an increased familial risk for MINOCA having a first-degree relative with MI-CAD, without an increased familial risk of MINOCA having a first-degree relative with MINOCA compared to controls. Age- and sex-adjusted hazard ratio (HR) for MICAD was 1.10 (95% confidence interval 1.03-1.18), and for MINOCA 0.99 (95% confidence interval 0.80-1.23). These results may indicate that genetic factors and shared environmental factors within a family leading to CAD are important also for MINOCA.

## Introduction

5-10% of all myocardial infarction (MI) present as a myocardial infarction with non-obstructive coronary arteries (MINOCA), defined as degree of stenosis of less than 50% on coronary angiography (1–3). It is increasingly recognized that a working diagnosis of MINOCA should be distinguished from a true diagnosis of MINOCA (4). The initial diagnosis of MINOCA should be considered a working diagnosis until other potential causes of the presentation are excluded, preferably by early cardiac magnetic resonance (CMR) imaging (5). A working diagnosis thus includes different underlying causes such as plaque rupture, spasm, dissection, thromboembolism, myocarditis, and takotsubo syndrome (5–8). Compared to myocardial infarction due to coronary artery disease (MI-CAD), patients with MINOCA are younger, more frequently women and have a lower prevalence of dyslipidemia, and diabetes mellitus (3,9–12). Others report that hypertension is more prevalent among MINOCA as compared with MI-CAD patients (3,12). In a Swedish register study, about 50% of the patients with MINOCA who had a reinfarction after their first event had progressed to MI-CAD (13). Apart from traditional CAD risk factors, a positive family history has been identified as an independent risk factor both for the development and recurrence of MI in patients with MI-CAD (14–18). This risk is proportional with the number of affected relatives and is stronger at young age (15,18,19). In two systematic reviews, self-reported family history of premature MI-CAD was similar between MINOCA and MI-CAD patients and more common compared with control subjects (3,10).

Even though familial risk is important for MI, the familial risk in patients with a working diagnosis MINOCA has not been investigated systematically before. Furthermore, previous studies on family history of MI have not distinguished between MINOCA and MI-CAD based on coronary angiographic findings. Therefore, our aim was to investigate the familial risk of a working diagnosis MINOCA in a nationwide cohort, with a secondary aim to investigate familial risk of MI-CAD among patients with MINOCA.

## Material and methods

### Study design

Nationwide register-based cohort study including MINOCA patients, and two different control groups identified from the Swedish coronary angiography and angioplasty register (SCAAR) based on a final diagnosis. This register is part of the Swedish web-system for enhancement and development of evidence-based care in heart disease evaluated according to recommended therapies (SWEDEHEART) register. All cohorts were defined by their diagnosis at the time of their first coronary angiography during the study period and only unique patients were investigated. All cases had a working diagnosis of MINOCA, defined by the absence of coronary artery stenosis of <50%, in SCAAR and a final diagnosis of MI in SWEDEHEART, and were labelled MINOCA in the context of this investigation. The first control cohort comprised patients with MI-CAD. They were defined by coronary artery stenosis ≥50% in SCAAR and a final diagnosis of MI in SWEDEHEART. The second control cohort comprised subjects who underwent coronary angiography for any indication e.g., chest pain, angina, valvular heart disease and cardiomyopathies but without MI in SWEDEHEART and had a non-obstructive coronary angiography defined by the absence of coronary artery stenosis of <50% in SCAAR. Among both MINOCA and control subjects without MI and obstructive coronary arteries, previous coronary artery bypass grafting (CABG) and percutaneous coronary intervention (PCI) were exclusion criteria as well as previous MI. For detailed participant selection view flow chart (Figure 1). The entire study period spanned from the start of 1995 until the end of 2020. All subjects in the cohorts were followed until a first degree relative experienced MINOCA or MI-CAD, or until censoring due to death or at the end of follow-up 31st December 2020, whichever occurred first.

**Figure 1.**
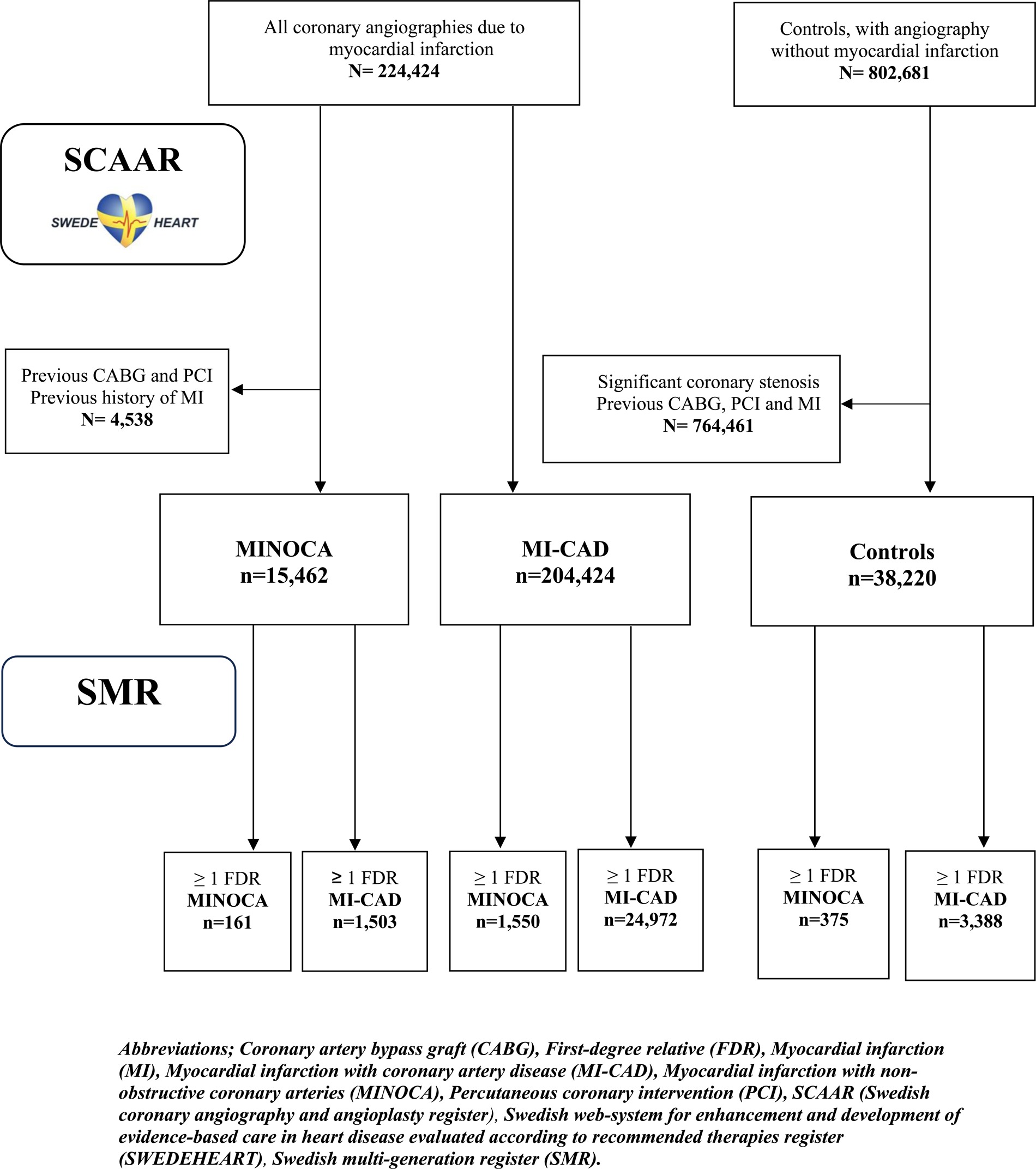
Flow chart of participant selection

### Ethical approval

Ethical permit for the study was granted by the Regional Ethical Review Board in Stockholm, Sweden (ethical permit number 2021-05506-02).

### Registers

SWEDEHEART is a national quality register that contains comprehensive information on various aspects of cardiac care in Sweden (20). It consists of several sub-divisions where SCAAR holds information of all coronary angiographies performed in Sweden. The register is regularly monitored with an accuracy of more than 90%. The diagnosis of MI is ascertained by cross-checking with the national patient register (20). Information on coronary angiographic findings was collected from SCAAR. Since 1995 all coronary angiographies have been included in SCAAR. Another subdivision of SWEDEHEART register is the Registry of Information and Knowledge about Swedish Heart Intensive Care Admission (RIKS-HIA) which includes detailed clinical information on previous CAD risk factors and comorbidities during cardiac care.

The Swedish multi-generation register (SMR) contains information on individuals that are either born or nationally registered in Sweden, from the years 1932 onwards and 1961, respectively. The register is managed by Statistics Sweden and contains information about connections between biological relatives, including parents, siblings, and children. The coverage rate is >95% on information of parents (21).

Data from SWEDEHEART were merged with data on first degree relatives from the SMR as well as information on hospital discharge diagnoses. The merging of information on an individual level between the two registries was made possible by using the unique personal identification number given all Swedish citizens.

### Statistical analysis

Baseline characteristics for the cohorts are presented as absolute numbers and proportions for categorical variables and means with standard deviation (SD) for continuous variables. Baseline data on index patients and relatives were collected from SWEDEHEART; Age, sex, previous MI and CABG, smoking status, previously diagnosed heart failure and stroke, hypertension, diabetes mellitus and hyperlipidemia where the latter was defined as ongoing treatment at the time of diagnosis as presented in the register. Smoking status was defined according to three categories: never, former, and current. Differences between two groups were tested using unpaired t-test for continuous variables and Fisher exact chi-square test for categorical data. Missing values are presented among covariates, and as these were less than 10%, (apart from body mass index), no imputations were made. We present baseline characteristics on all patients (Table 1), on complete cases (Supplementary Table 1B) and on relatives (Table 2) and index patients to relatives (Supplementary Table 2B).

**Table 1.**
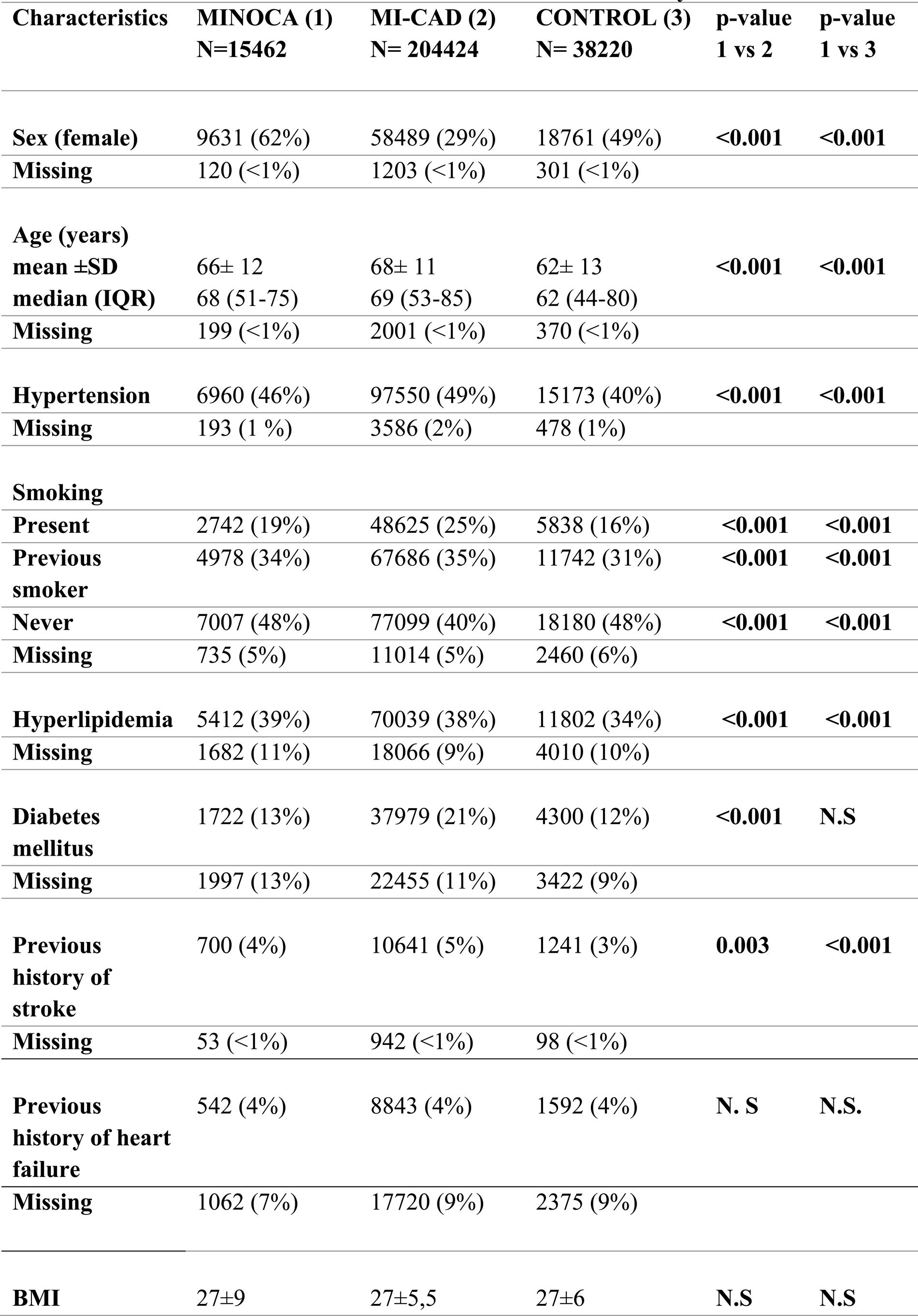
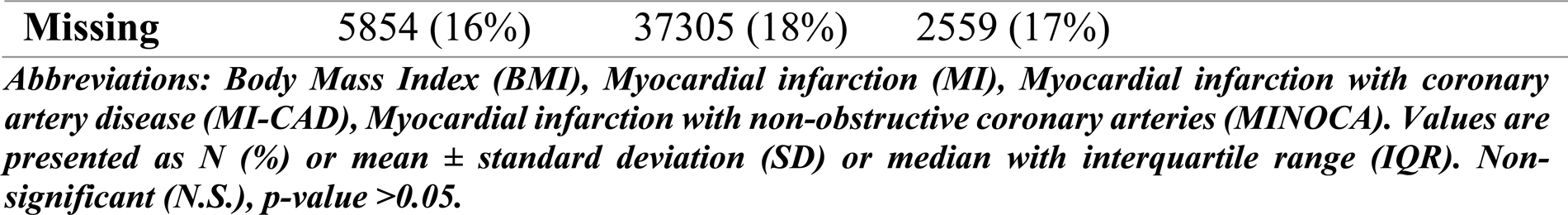
Baseline characteristics for study participants with MINOCA, MI-CAD and controls without MI and without obstructive coronary arteries.

| **Characteristics** | **MINOCA (1)**<br>**N=15462** | **MI-CAD (2)**<br>**N= 204424** | **CONTROL (3)**<br>**N= 38220** | **p-value**<br>**1 vs 2** | **p-value**<br>**1 vs 3** |
| --- | --- | --- | --- | --- | --- |
| **Sex (female)** | 9631 (62%) | 58489 (29%) | 18761 (49%) | **<0.001** | **<0.001** |
| **Missing** | 120 (<1%) | 1203 (<1%) | 301 (<1%) |  |  |
| **Age (years)** |  |  |  |  |  |
| **mean ±SD** | 66± 12 | 68± 11 | 62± 13 | **<0.001** | **<0.001** |
| **median (IQR)** | 68 (51-75) | 69 (53-85) | 62 (44-80) |  |  |
| **Missing** | 199 (<1%) | 2001 (<1%) | 370 (<1%) |  |  |
| **Hypertension** | 6960 (46%) | 97550 (49%) | 15173 (40%) | **<0.001** | **<0.001** |
| **Missing** | 193 (1 %) | 3586 (2%) | 478 (1%) |  |  |
| **Smoking** |  |  |  |  |  |
| **Present** | 2742 (19%) | 48625 (25%) | 5838 (16%) | **<0.001** | **<0.001** |
| **Previous smoker** | 4978 (34%) | 67686 (35%) | 11742 (31%) | **<0.001** | **<0.001** |
| **Never** | 7007 (48%) | 77099 (40%) | 18180 (48%) | **<0.001** | **<0.001** |
| **Missing** | 735 (5%) | 11014 (5%) | 2460 (6%) |  |  |
| **Hyperlipidemia** | 5412 (39%) | 70039 (38%) | 11802 (34%) | **<0.001** | **<0.001** |
| **Missing** | 1682 (11%) | 18066 (9%) | 4010 (10%) |  |  |
| **Diabetes mellitus** | 1722 (13%) | 37979 (21%) | 4300 (12%) | **<0.001** | **N.S** |
| **Missing** | 1997 (13%) | 22455 (11%) | 3422 (9%) |  |  |
| **Previous history of stroke** | 700 (4%) | 10641 (5%) | 1241 (3%) | **0.003** | **<0.001** |
| **Missing** | 53 (<1%) | 942 (<1%) | 98 (<1%) |  |  |
| **Previous history of heart failure** | 542 (4%) | 8843 (4%) | 1592 (4%) | **N. S** | **N.S.** |
| **Missing** | 1062 (7%) | 17720 (9%) | 2375 (9%) |  |  |
| **BMI** | 27±9 | 27±5,5 | 27±6 | **N.S** | **N.S** |

|  |  |  |  |
| --- | --- | --- | --- |
| **Missing** | 5854 (16%) | 37305 (18%) | 2559 (17%) |

*Abbreviations: Body Mass Index (BMI), Myocardial infarction (MI), Myocardial infarction with coronary artery disease (MI-CAD), Myocardial infarction with non-obstructive coronary arteries (MINOCA). Values are presented as N (%) or mean ± standard deviation (SD) or median with interquartile range (IQR). Non-significant (N.S.), p-value >0.05.*

**Table 2.**
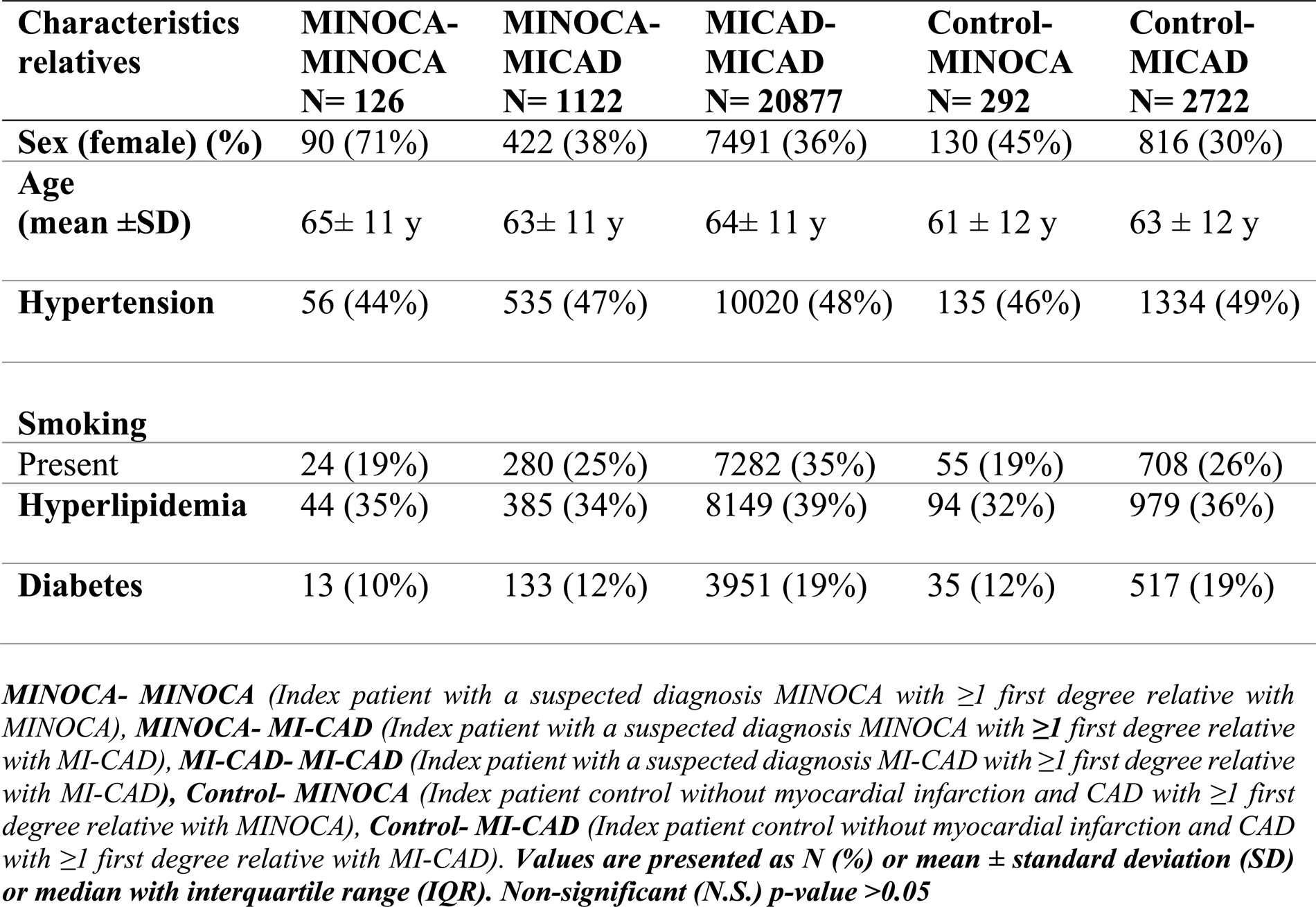
Baseline characteristics of first-degree relatives with MINOCA and MICAD.

| **Characteristics**<br>**relatives** | **MINOCA-**<br>**MINOCA**<br>**N= 126** | **MINOCA-**<br>**MICAD**<br>**N= 1122** | **MICAD-**<br>**MICAD**<br>**N= 20877** | **Control-**<br>**MINOCA**<br>**N= 292** | **Control-**<br>**MICAD**<br>**N= 2722** |
| --- | --- | --- | --- | --- | --- |
| **Sex (female) (%)** | 90 (71%) | 422 (38%) | 7491 (36%) | 130 (45%) | 816 (30%) |
| **Age**<br>**(mean ±SD)** | 65± 11 y | 63± 11 y | 64± 11 y | 61 ± 12 y | 63 ± 12 y |
| **Hypertension** | 56 (44%) | 535 (47%) | 10020 (48%) | 135 (46%) | 1334 (49%) |
| **Smoking** |  |  |  |  |  |
| Present | 24 (19%) | 280 (25%) | 7282 (35%) | 55 (19%) | 708 (26%) |
| **Hyperlipidemia** | 44 (35%) | 385 (34%) | 8149 (39%) | 94 (32%) | 979 (36%) |
| **Diabetes** | 13 (10%) | 133 (12%) | 3951 (19%) | 35 (12%) | 517 (19%) |

**MINOCA- MINOCA** (Index patient with a suspected diagnosis MINOCA with $\geq 1$ first degree relative with MINOCA), **MINOCA- MI-CAD** (Index patient with a suspected diagnosis MINOCA with $\geq 1$ first degree relative with MI-CAD), **MI-CAD- MI-CAD** (Index patient with a suspected diagnosis MI-CAD with $\geq 1$ first degree relative with MI-CAD), **Control- MINOCA** (Index patient control without myocardial infarction and CAD with $\geq 1$ first degree relative with MINOCA), **Control- MI-CAD** (Index patient control without myocardial infarction and CAD with $\geq 1$ first degree relative with MI-CAD). **Values are presented as N (%) or mean ± standard deviation (SD) or median with interquartile range (IQR). Non-significant (N.S.) p-value >0.05**

Cox proportional hazard regression models were used to estimate the risk of having a first-degree relative experiencing MINOCA or MI-CAD during the follow-up compared to control subjects with non-obstructive coronary arteries. As controls without CAD and MICAD patients differed significantly in age and sex from MINOCA patients, our model was adjusted for sex and age. In a multivariable analysis adjustment was also made for smoking and the following comorbidities: hypertension, hyperlipidemia and diabetes mellitus.

In the current study, first-degree relatives were defined as parents, children or “full siblings” whereas half-siblings were not included. We defined time at risk for an event (a first-degree relative experiencing MINOCA or MI-CAD) as time from a diagnosis of MINOCA, MI-CAD or a non-obstructive coronary angiography without MI to a MINOCA or MI-CAD diagnosis in a first degree relative, death or to the end of follow-up (31 December 2020), whichever occurred first. Age was included as a categorical variable, stratified in three age-groups with estimated equal risk of exposure to disease in the model. Subgroup analyses were performed for women and men and for patients with premature MINOCA and MI-CAD separately.

Premature MINOCA was defined as receiving the diagnosis at age 65 years or younger. Premature MI-CAD was defined as receiving a myocardial infarction diagnosis at age 65 or younger in women, and 55 years or younger in men (22). All analyses were performed with R V.3.4. and SPSS 27 (IBM, Armonk, New York).

## Results

A total of 15,462 MINOCA patients, 204,424 MI-CAD patients, together with 38,220 control subjects without MI with a non-obstructive coronary angiography were included into the study (Figure 1). MINOCA and MI-CAD patients were older than control subjects without MI with a non-obstructive coronary angiography. MI-CAD patients were more often males and had more comorbidities such as hypertension, diabetes mellitus, and smoking, compared to MINOCA patients and control subjects. MINOCA patients were younger and more often women compared to MI-CAD patients and had less CAD risk factors compared to MI-CAD except for hyperlipidemia. In contrast, MINOCA patients had more CAD risk factors compared to control subjects apart from diabetes mellitus (Table 1) (Supplement Table 1B). Relatives with MINOCA and MI-CAD were slightly younger than each cohort in total (Table 2).

### Familial risk of MINOCA

During a mean follow-up of 8.1 ± 4.2 years, MINOCA occurred in a first-degree relative to 161 out of 15,462 patients initially diagnosed with MINOCA (1.0 %). Control subjects without MI with non-obstructive coronary arteries were followed for 7.4 ±4.4 years and had a first-degree relative with MINOCA in 1.0 % of the cases. The age- and sex-adjusted HR for familial risk of MINOCA for 15,023 MINOCA cases compared to control subjects without MI with a non-obstructive coronary angiography was HR 0.99 (95% CI 0.80-1.23). A total of 11,991 MINOCA patients had complete information on event date and all covariates and were included in an adjusted multivariable Cox regression analysis. Of these, information on risk exposures and all covariates was available in 126 MINOCA-relatives, of which 90 (70.6%) were women. HR adjusted for all covariates was 0.97 (95% CI 0.79-1.21). In subgroup analyses results were similar among females and males with MINOCA, and for MINOCA-patients 65 years or younger, adjusted for all covariates (Figure 2).

**Figure 2.**
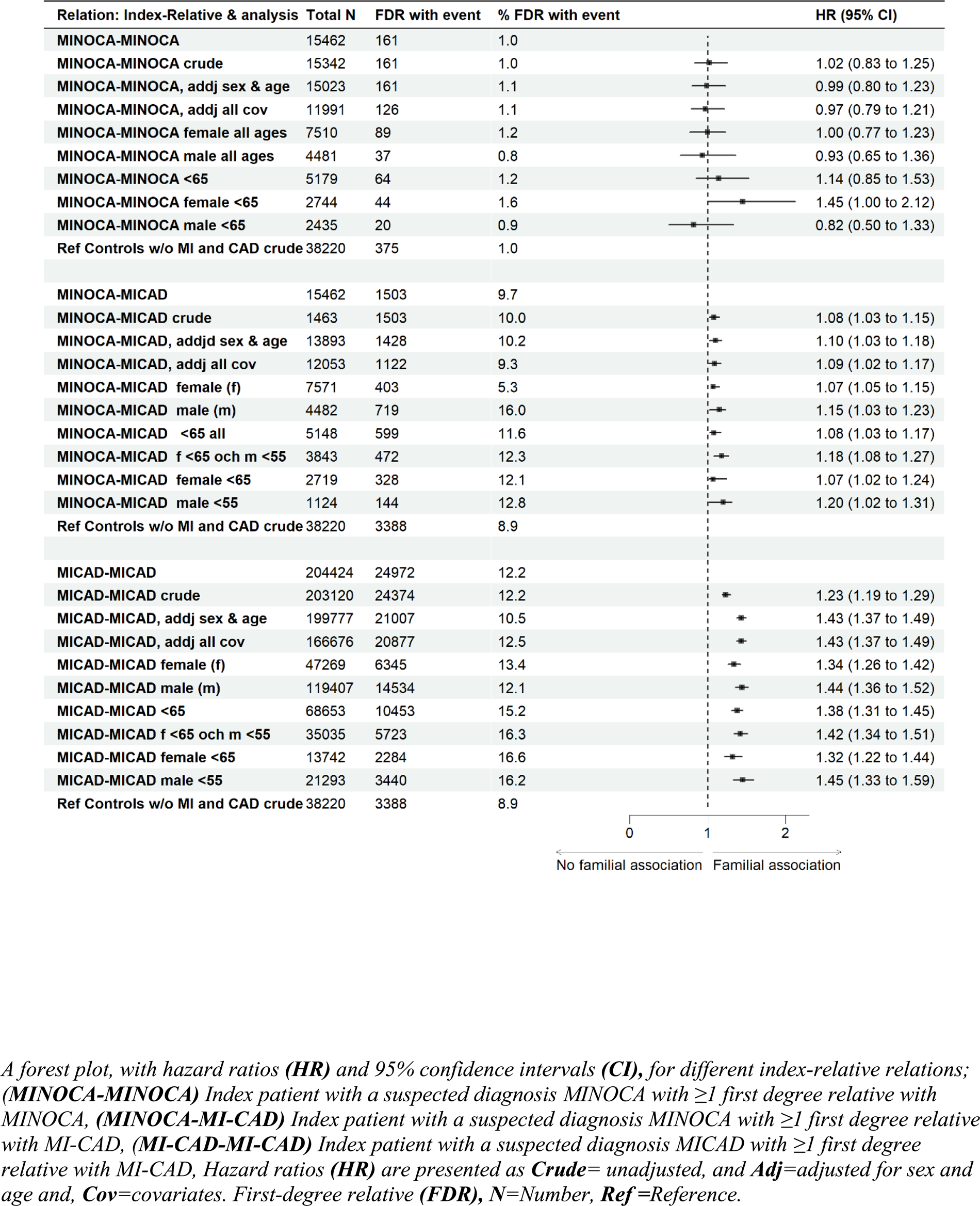
Forest plot and table of family history of MINOCA and MI-CAD for individuals with MINOCA and of family history of MI-CAD for individuals with MI-CAD

### Familial risk of MI-CAD

During a mean follow-up period of 8.2 ±4.4 years, a diagnosis of MI-CAD occurred in 1,503 first-degree relatives out of 15,462 patients diagnosed with MINOCA (9.7%). Control subjects followed for 7.6 ±4.0 years had a first-degree relative with a MI-CAD in 8.9% of the cases. The age- and sex-adjusted HR for familial risk of MI-CAD among 13,893 MINOCA cases compared to control subjects without MI with a non-obstructive coronary angiography was 1.10 (95% CI 1.03-1.18). Among MI-CAD relatives, 1122 had available information on risk exposure and relevant covariates of which 422 (37.6%) were women. HR adjusted for all covariates was 1.09 (95% CI 1.02-1.17). In subgroup analyses the results were similar among females and males with MI-CAD, and in premature MI-CAD,.(Figure 2).

During a mean follow-up of 8.5 ±4.8 years, a diagnosis of MI-CAD occurred in a first-degree relative of 24,972 MI-CAD out of 204,424 patients (12.2%) in comparison to 8.9 % of the control subjects. The age- and sex-adjusted HR for familial risk MI-CAD of 199,777 MI-CAD cases compared to control subjects without MI with a non-obstructive coronary angiography was 1.43 (95% CI 1.37-1.49). 20,877 relatives had information on risk exposure and all covariates of which 7491 (36%) MI-CAD patients were females. HR adjusted for all covariates was 1.43 (95% CI 1.37-1.49). The HR tended to be higher for men than women. In a subgroup analysis according to sex HR for men was 1.44 (95% CI 1.36-1.52) and 1.34 (95% CI 1.26-1.42) for women when adjusted for all covariates. In a subgroup analysis of premature MI-CAD, HR remained unchanged (Figure 2).

## Discussion

This is the first study that examines familial risk of MINOCA and MI-CAD based on coronary angiographic findings, including data from a nationwide sample including over 250,000 patients with a mean follow-up of 8 years. A familial risk of a working diagnosis of MINOCA was only found among 1% of all MINOCA patients and was not elevated when compared to control subjects without MI with a non-obstructive coronary angiography. In contrast, an adjusted 10% familial risk for MI-CAD was apparent among patients with a working diagnosis MINOCA which was higher compared to control subjects. This suggests that a significant proportion of MINOCA patients share underlying mechanisms with MI-CAD.

### Familial risk of MINOCA

To the best of our knowledge, no study has investigated family history in patients with MINOCA combining high quality registers with data on coronary angiography results and heritability. Of note, the present study did only include patients with an ascertained diagnosis of MI. However, we cannot exclude that some patients had spontaneous coronary artery dissection (SCAD) or takotsubo syndrome. Patients with SCAD could have been categorized either as MINOCA or MICAD. Takotsubo has been excluded from a definite diagnosis of MINOCA after the introduction of the 4^th^ universal definition of MI (23). In SCAD, genome-wide association studies have identified genetic risk variants, however studies have been small and results conflicting (24). In takotsubo syndrome, reports of familial cases, high recurrence rates and case reports of patients with rare genetic syndromes supports a familial component (25–27). The lack of evidence of a family history of a MINOCA in our study is not proof of absence of a family history of MINOCA but could be a result of limitations in the study design, including sample size and selection. The absolute number of MINOCA patients with first degree relatives with MINOCA was low, making further exploratory sub-group analyses difficult. Lack of evidence of a positive family history of MINOCA highlights the importance of additional studies of MINOCA and related groups of patients such as SCAD and takotsubo syndrome(28).

### Familial risk of MI-CAD

MINOCA patients were more often found to have a familial risk of MI-CAD when compared to control subjects without MI with a non-obstructive coronary angiography. The association was significant, however weaker than for MI-CAD patients. This could indicate that similar mechanisms are of importance for disease development both for MINOCA and for MI-CAD. The weaker association indicates that certain sub-groups without family history of MI-CAD within MINOCA are prevalent when compared to patients with MI-CAD. Results are in line with a previous meta-analysis by Pasupathy et al. including data from six separate studies on MINOCA (10). These studies rely on self-reported data of family history with its obvious limitation of recall-bias and have limitations on different definitions of family history. The close relationship between MINOCA and MI-CAD is highlighted by the arbitrary definition of a significant stenosis ≥50% that in part could explain the results. The lack of further coronary imaging results such as intravascular ultrasound or optical coherence tomography (OCT) and the fact that silent coronary artery stenosis is common could lead to misclassification. This is illustrated by a population-based Swedish study, 5.2% of the middle-aged Swedish population had significant coronary stenosis without MI (29). There are several studies supporting similar mechanisms behind MINOCA and MI-CAD. One Swedish register study examined the recurrence of MI in MINOCA. The recurrence rate was shown to be around 6% and about 50% of the patients with a recurrent MI had progressed to MI-CAD. Furthermore, Reynolds and coworkers (30) showed that almost half of a group of female MINOCA patients had a culprit lesion as determined by OCT. Taken together, the results from this study and others indicate that CAD could be the underlying cause in a significant proportion of MINOCA cases.

The results for patients with MI-CAD are in concordance with previous studies on family history of MI. It is well established that heritability plays an important role in the development of MI, which is supported by twin studies, case-control/cohort studies of family history and studies on single-nucleotide polymorphisms (SNP) associated with MI (31). Today over 300 SNPs associated with cardiovascular disease have been identified. In our study MI-CAD patients had a familial risk of myocardial infarction with a HR of 1.45 when adjusting for known CAD risk factors. This is similar to previous studies such as the INTERHEART where a family history of MI was associated with MI with an OR of 1.84 for paternal family history and OR 1.72 for maternal history and the Framingham Offspring study with a family history among siblings was associated with CVD with an OR adjusted for CVD risk factors of 1.45. Many of the previous family studies have relied on self-reported data, which is a limitation, instead of objective register data. Taken together, our results support previous studies on family history of MI-CAD as an independent risk factor for MI.

### Strengths and limitations

The major strength of our study lies in the large sample size and unbiased selection, including all MIs that underwent a coronary angiography in Sweden between 1995-2020, which increases the reliability and generalizability of the study. Another strength is the diagnostic confirmation of a diagnosis of MINOCA and MI-CAD by coronary angiographic findings. Furthermore, the register-based design offers advantages when compared to self-reported family history, since it minimizes recall bias. This design has been used successfully in various settings (19, 31). Finally, SCAAR and SMR have been shown to have both a high coverage and data validity (20, 21). The fact that we could replicate previous studies on MI-CAD confirms the study design.

The study has several limitations; One limitation is that the use of cases and controls from the SCAAR register might introduce a selection bias towards patients investigated during the last decade when the use of coronary angiography has been a common diagnostic tool. Another limitation is uncertainty in diagnosis as in all register studies. Another limitation is that we did not have information on all family members. We lack information on siblings and parents with MI who did not perform a coronary angiography during the study period and if the total numbers of relatives were different between cases and controls. Furthermore, despite the considerable number of included individuals in the study, the total number of first-degree relatives with a working diagnosis MINOCA was low, presumably the study results depended mainly on siblings due to the study period of 25 years. Furthermore, only full siblings, with the same mother and father, were included in the study, thus there might have been an underreporting on family history. Other limitations include the absence of certain baseline characteristics, such as socioeconomic and environmental factors, which are important risk factors for myocardial infarction. Hence, important known and unknown confounders could be missing. Finally, the choice of control subjects from a group without MI with non-obstructive coronary arteries does not represent a healthy population since they had an indication for their coronary angiography. The differences in familial risk might have been more pronounced with a healthy control group.

### Clinical perspectives

The recent European guidelines recommends comprehensive risk assessment and systematic screening for cardiovascular disease (CVD) in subjects with a family history of premature CVD, defined as 50-65 years old (32). Evidence of a familial risk for MI-CAD among patients with MINOCA is important when choosing secondary prevention and treatment. In the future, more focus will be on personalized primary prevention and individualized treatment thus elucidating common mechanisms for MINOCA and MI-CAD are of importance for future treatment strategies.

## Conclusion

No increased familial risk of MINOCA was observed for MINOCA-patients whereas there was an increased familial risk for MI-CAD when compared to control subjects with non-obstructive coronary arteries. These results could indicate that genetic factors and shared environmental factors within a family leading to CAD are important also for MINOCA, thus MI-CAD and MINOCA may share underlying mechanisms.

## Data Availability

Data will be available on demand

## List of abbreviations

CABG: coronary artery bypass grafting
CAD: coronary artery disease
MINOCA: myocardial infarction with non-obstructive coronary arteries
MI: myocardial infarction
MI-CAD: myocardial infarction with coronary artery disease
PCI: percutaneous coronary intervention
SCAAR: Swedish coronary angiography and angioplasty registry
SMR: Swedish multi-generation register
SWEDEHEART: Swedish web-system for enhancement and development of evidence-based care in heart disease evaluated according to recommended therapies register.

## Funding

Swedish Heart and Lung Foundation

## Disclosures

None

## Acknowledgements

None

## Notes

### Competing Interest Statement

The authors have declared no competing interest.

### Author Declarations

Ethical permit (Regional Ethical Review Board in Stockholm, Sweden) no. 2018/23-32

